# The prevalence of antibodies to SARS-CoV-2 in asymptomatic healthcare workers with intensive exposure to COVID-19

**DOI:** 10.1101/2020.05.28.20110767

**Authors:** Shue Xiong, Chunxia Guo, Ulf Dittmer, Xin Zheng, Baoju Wang

## Abstract

The prevalence of asymptomatic SARS-CoV-2 infection in healthcare workers with intensive exposure to COVID-19 is unclear. In this study, we investigated the seroprevalence of SARS-CoV-2 in 797 asymptomatic healthcare workers with intensive exposure to COVID-19 patients in Wuhan, China. Positive IgG was detected from 35 asymptomatic healthcare workers, and the prevalence of antibodies to SARS-CoV-2 in asymptomatic healthcare workers was 4.39% (35/797). None of them developed COVID-19 until May 15. 33 of them have performed at least one chest CT scan showing no viral pneumonia features, and 16 have finished at least one-time SARS-CoV-2 RNA detection with negative results. When contacting with the patients, 15 of them dressed with full personal protective equipment (PPE), and 16 worn N95 mask and gown. To the best of our knowledge, this is the first investigation reported that the seroprevalence of SARS-CoV-2 was 4.39% in asymptomatic healthcare workers with applied PPE in a high epidemic area, which may provide useful information of estimating asymptomatic infection rate in general population.

## Introduction

The 2019 novel coronavirus disease (COVID-19) caused by severe respiratory syndrome coronavirus 2 (SARS-CoV-2) is pandemic worldwide^1-3^. Asymptomatic SARS-CoV-2 infection has been concerned to be risk for spreading virus^4^, while the prevalence of asymptomatic SARS-CoV-2 infection in healthcare workers with intensive exposure to COVID-19 is unclear. In this study, we investigated the seroprevalence of SARS-CoV-2 in asymptomatic healthcare workers with intensive exposure to COVID-19 patients in Wuhan, China.

## Methods

The epidemiological information was collected by trained investigators from 797 asymptomatic healthcare workers (705 female) in Wuhan. Blood was drawn from February 12, 2020, to March 17, 2020. Serological tests of IgM and IgG antibodies against SARS-CoV-2 were performed using an ELISA assay. The interval from the first time of exposure to date of antibody detection was 37-72 days. This study was approved by medical ethics committee of Union Hospital, Huazhong University of Science and Technology (0056).

## Results

Positive IgG was detected from 35 asymptomatic healthcare workers, and the prevalence of antibodies to SARS-CoV-2 in asymptomatic healthcare workers was 4.39% (35/797). The epidemiological information and test results were shown in table 1. None of them developed COVID-19 until May 15. 33 of them have performed at least one chest CT scan showing no viral pneumonia features, and 16 have finished at least once SARS-CoV-2 RNA detection with negative results. All participants were self-isolated during and after finishing their duties before antibody testing. None of their family members was infected with SARS-CoV-2. The median age was 31 years (range 23-53), and all were female. None of them had history of smoking, drinking, chronic diseases and medication. Among these 35 cases, one is physician; one is radiology technician, whereas 33 are nursing staff. Twenty-six of them reported to have close contact with confirmed COVID-19 patients. Four of them had no direct exposure to patients and 5 workers were uncertain of their exposure time. When contacting with the patients, 15 of them dressed with full personal protective equipment (PPE), and 16 worn N95 mask and gown. Seventeen nurses performed high-risk operations, including sputum suction, ventilator operation, and sampling.

**Table 1.**
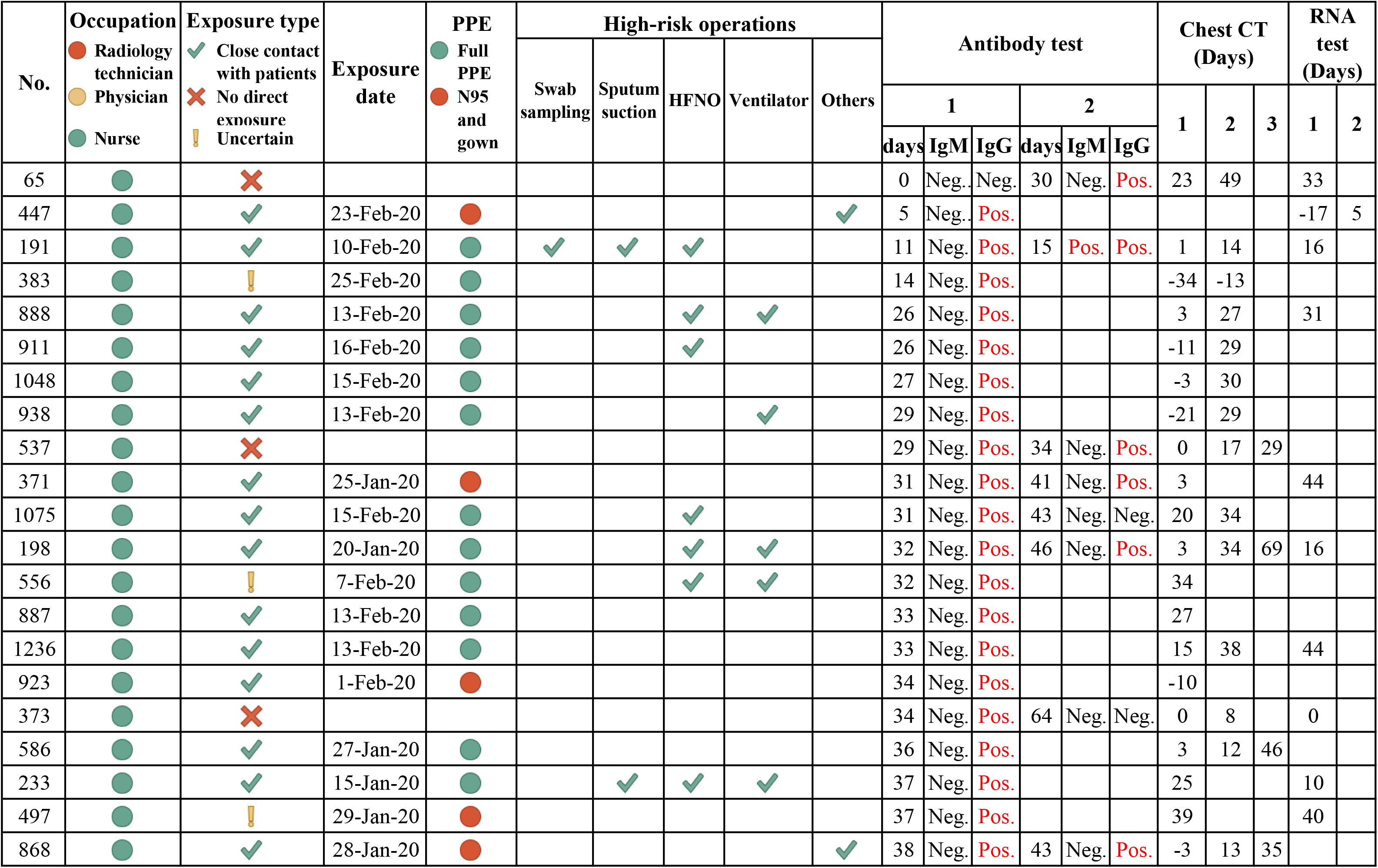
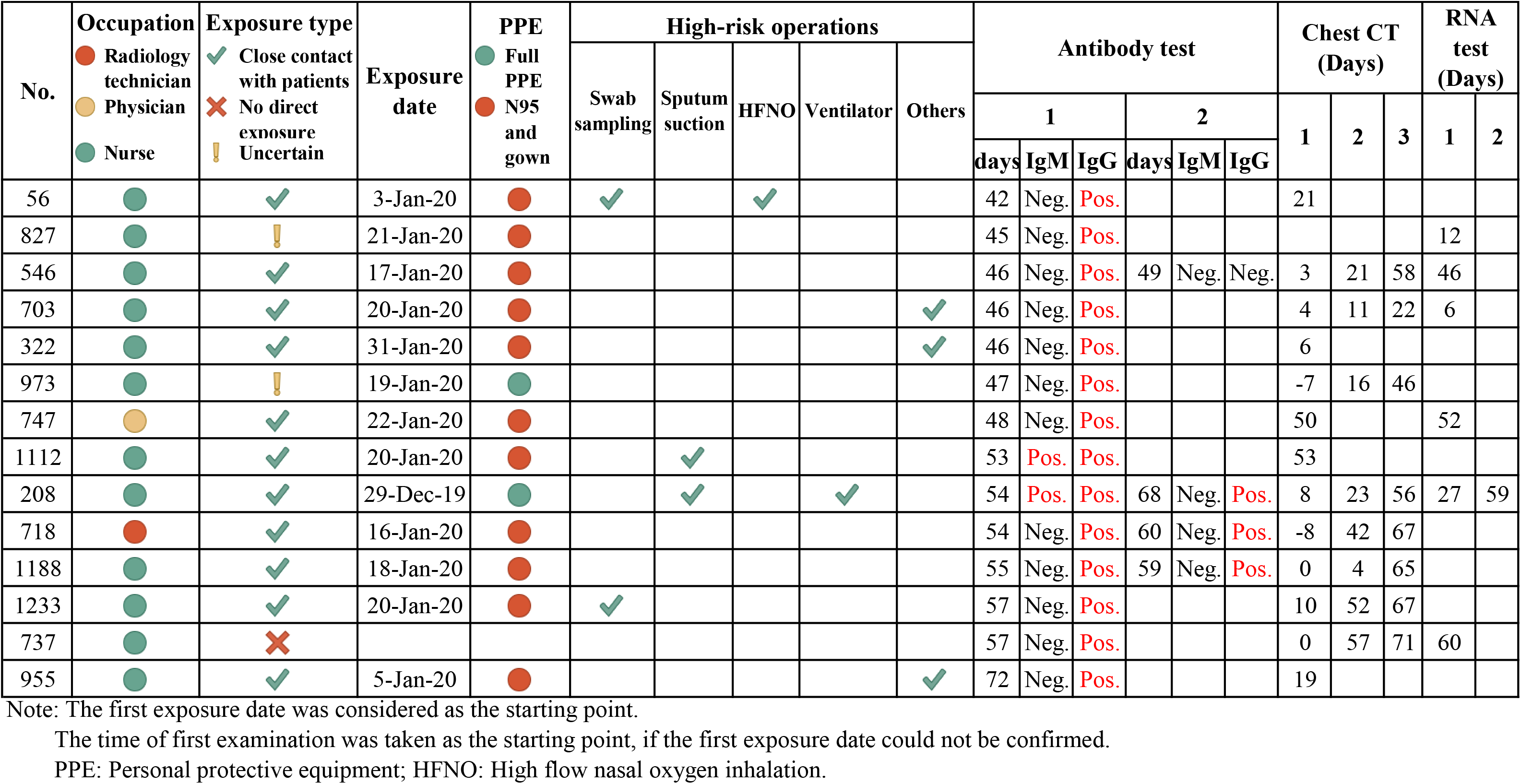
The detailed information of 35 healthcare workers with asymptomatic SARS-CoV-2 infection.

## Discussion

The healthcare workers are at higher risk of SARS-CoV-2 infection due to their high frequency of exposing to patients and performing high-risk operations. To the best of our knowledge, this is the first investigation reported that the seroprevalence of SARS-CoV-2 was 4.39% in asymptomatic healthcare workers with applied PPE in a high epidemic area. These asymptomatic carriages were young, less likely to have comorbid diseases and did not develop COVID-19. The extent of seroconversion in asymptomatic healthcare workers may provide useful information of estimating asymptomatic infection rate in general population.

There is a great concern about the role of asymptomatic healthcare workers in the transmission of SARS-CoV-2 in hospital and community. It is not realistic to closely monitor the medical staff without any symptoms because the SARS-CoV-2 RNA may exist in a narrow window. Application of PPE when medical staff performed operations and keeping them self-isolated during and after finishing duties in hospital are very important measures for preventing transmission.

## Data Availability

The raw/processed data required to reproduce these findings cannot be shared at this time as the data also forms part of an ongoing study.

## Author Contributions

Dr S.E.X., C.X.G., and B.J.W. had full access to all of the data in the study and takes responsibility for the integrity of the data and the accuracy of the data analysis. Joint first authors are S.E.X. and C.X.G.

Study concept and design: X. Z., and B.J.W.

Acquisition, analysis, or interpretation of data: S.E.X, C.X.G, and B.J.W.

Drafting of the manuscript: S.E.X., X.Z., and B.J.W.

Critical revision of the manuscript for important intellectual content: U.D.

## Acknowledgment

This study was partly supported by the National Science and Technology Major Project for Infectious Diseases of China (2017ZX10304402-002-005, 2018ZX10723203, 2018ZX10302206), HUST COVID-19 Rapid Response Call (2020kfyXGYJ046, 2020kfyXGYJ016), and the Medical Faculty of the University of Duisburg-Essen and Stiftung Universitaetsmedizin, University Hospital Essen, Germany. The funders had no role in the design and conduct of the study; collection, management, analysis, and interpretation of the data; preparation, review, or approval of the manuscript; and decision to submit the manuscript for publication.

## Patient consent

We obtained written informed consent from the participants and this study was approved by medical ethics committee of Union Hospital, Huazhong University of Science and Technology (0056).

## Additional Contributions

We thank Dr. Yanfang Zhang (Wuhan Institute of Virology, Chinese Academy of Sciences), Weixian Wang and Min Liao (Department of Infectious Diseases, Union Hospital, Tongji Medical College, Huazhong University of Science and Technology) for technical assistance, and Prof. Dr. Mengji Lu (Institute for Virology, University Hospital of Essen, University of Duisburg-Essen) for manuscript revision.

